# Differential COVID-19 infection rates in children, adults, and elderly: evidence from 38 pre-vaccination national seroprevalence studies

**DOI:** 10.1101/2022.06.28.22277034

**Authors:** Cathrine Axfors, Angelo Maria Pezzullo, Despina G. Contopoulos-Ioannidis, Alexandre Apostolatos, John P.A. Ioannidis

## Abstract

**Background:** COVID-19 exhibits a steep age gradient of infection fatality rate. There has been debate about whether extra protection of elderly and other vulnerable individuals (precision shielding) is feasible, and, if so, to what extent.

**Methods:** We used systematically retrieved data from national seroprevalence studies conducted in the pre-vaccination era. Studies were identified through SeroTracker and PubMed searches (last update May 17, 2022). Studies were eligible if they targeted representative general populations without high risk of bias. Seroprevalence estimates were noted for children, non-elderly adults, and elderly adults, using cut-offs of 20, and 60 years (or as close to these ages, if they were not available).

**Results:** Thirty-eight national seroprevalence studies from 36 different countries were included in the analysis. 26/38 also included pediatric populations. 25/38 studies were from high-income countries. The median ratio of seroprevalence in the elderly versus non-elderly adults (or non-elderly in general, if pediatric and adult population data were not offered separately) was 0.90-0.95 in different analyses with large variability across studies. In 5 studies (all of them in high-income countries), there was significant protection of the elderly with ratio <0.40. The median was 0.83 in high-income countries and 1.02 in other countries. The median ratio of seroprevalence in children versus adults was 0.89 and only one study showed a significant ratio of <0.40.

**Conclusion:** Precision shielding of elderly community-dwelling populations before the availability of vaccines was feasible in some high-income countries, but most countries failed to achieve any substantial focused protection of this age group.

**summary:** 38 COVID-19 nationally representative seroprevalence studies conducted before vaccination campaigns were systematically identified. Median seroprevalence ratio in elderly versus non-elderly adults was 0.90-0.95, indicating no generally achieved precision shielding of elderly. In 5 studies, substantial protection (ratio <0.40) was observed.

## INTRODUCTION

Coronavirus Disease 2019 (COVID-19) is characterized by a steep age-gradient in risk of serious disease and death^1-3^. Death risk after infection increases ∼3-fold per 10-year increment, thus differing >1,000-fold between pediatric and geriatric populations. The total fatalities footprint of a pandemic with such strong risk stratification is expected to depend on how effectively high-risk, vulnerable individuals are protected from infection.^4^ This applies regardless of whether other effective interventions are used, such as vaccines. However, it is particularly important for the prevaccination period.

The ability to protect more aggressively the elderly and other vulnerable individuals (aka, precision shielding) has been contested^5,6^. For a widely circulating virus, it may be difficult to effectively shield only high-risk individuals. In fact, nursing home residents, a particularly high-risk group of elderly people, were even disproportionately more frequently infected early in the pandemic^7-11^. Infections were massively spread in such facilities, as testified by high death toll and high seroprevalence rates in their populations^9-13^. However, the question of age-stratified precision shielding remains open for community-dwelling populations. It is possible that infection rates varied in different age groups. Perhaps community-dwelling elderly might have been less mobile and less exposed than other adults. Conversely, at the other end of the age pyramid, children and adolescents may have had lower infection rates, given the widely implemented school closures.

Insights on the relative infection rates across age strata can be obtained from seroprevalence studies. Hundreds of such studies have been conducted to-date.^14^ However, such surveys are also susceptible to numerous biases.^15^ Here, to answer the question of whether age-specific precision shielding was achieved in the pre-vaccination period, we used data from comprehensive, national seroprevalence studies without high risk of bias.

## METHODS

The study was pre-registered as part of a broader ongoing project on COVID-19 seroprevalence and infection fatality (protocol: https://osf.io/xvupr). Protocol amendments/clarifications appear in Supplementary Table 1.

### Search strategy and eligibility criteria

We identified seroprevalence studies (articles, official reports, or preprints) in the live systematic review SeroTracker^14-16^. We also performed PubMed searches using the string “seroprevalence AND (national OR stratified) AND COVID-19” to identify potentially eligible studies that were recently published and may not have been indexed in SeroTracker yet. The searches were performed initially on February 8, 2022 and upon completion of the data extraction, we updated the searches on May 17, 2022 to examine if any additional studies had become available.

We included studies on SARS-CoV-2 seroprevalence that met the following criteria: sampled any number of participants in a national representative sample; sampling was completed by February 28, 2021; adults (>=21 years old) were included, regardless of whether children and/or adolescents were included or not; provided an estimate of seroprevalence for non-elderly people (preferably for <70 years and/or <60 years, but any cut-off between 60 and 70 years was acceptable); and explicitly aimed to generate samples reflecting the general population at a national level, excluding studies focusing on patient cohorts (such as residual clinical samples), blood donors, workers (healthcare or other), and insurance applicants as well as any other study where the examined population might have had lower or higher risk than the general population. While studies of blood donors, residual clinical samples and non-healthcare workers usually tend to give on average seroprevalence estimates similar to those of explicitly sampled general populations^15^, their results may occasionally be more biased and the representation and bias may be particularly affected in the elderly strata. In SeroTracker, only studies in the categories of “Household and community samples” and “Multiple general populations” without high risk of bias (reported by the SeroTracker team using the Joanna Briggs Institute Critical Appraisal Tool for Prevalence Studies) were considered for further scrutiny. Similar criteria were applied to any additional PubMed-retrieved studies. Furthermore, similar to previous work^17^, for studies done in the USA, only those that have adjusted the seroprevalence estimates for race/ethnicity were retained, since this factor is known to associate strongly with the risk of SARS-CoV-2 infection^18^.

For studies with several sampled (sub)regions of a country, we accepted those where the sampling locations were dispersed across the country so as to form a reasonable representation of the entire country. We excluded studies where the sampling locations were potentially biased towards higher or lower seroprevalence than the general population. For example, studies were excluded when only urban populations or when only rural populations were sampled; or when locations were selected because they were hard hit (e.g. had many deaths or many cases) or were lightly hit (e.g. had no detected cases or fewer than average detected cases). Conversely, studies composed of regional units were eligible if they included both high and low risk sampling units, aiming for a total sample that is fairly representative of the national general population.

We excluded seroprevalence studies where crude overall seroprevalence in the population was less than 1-test specificity and/or the 95% confidence interval of the seroprevalence went to 0%, since the uncertainty on seroprevalence for them is very large.

We also excluded studies that included in their sampling only pediatric populations without any adults 21 years or older. Studies that used in their sampling a lower age boundary that excluded children and/or adolescents and/or some young adults (e.g. >5, or >18, or >25 years) were included. Studies were included regardless of whether any upper age boundary was used in their sampling.

To avoid any substantive impact of vaccination, we only considered seroprevalence studies where the sampling had been completed by the end of February 2021 and at least 90% of the samples had been collected before end of January 2021.

### Extracted information

Data extraction for eligible articles was performed in duplicate by two authors independently and disagreements were discussed. In cases of persistent disagreements, a third author (JPAI) was the arbitrator.

We extracted from all eligible seroprevalence studies their information on country, dates of sample collection, overall sample size (number tested) and sample size in pediatric, non-elderly adults, and elderly populations, and types of antibody measured (immunoglobulin G (IgG), IgM, IgA). We also extracted the estimated unadjusted seroprevalence (positive samples divided by all samples tested), and the most fully adjusted seroprevalence in children, non-elderly adults, and elderly. We also noted the factors that the authors considered for adjustment in the most fully adjusted calculations. Whenever there were multiple different time points when seroprevalence was assessed in a given study, we selected the time point that gives the highest overall seroprevalence estimate and when there was a tie, we chose the earliest time point.

The groups of children (including adolescents), non-elderly adults, and elderly were defined according to preferred age cut-offs of 20 years and 60 years, therefore ideally these groups referred to 0-20 years, 21-60 years, and >60 years, respectively. For separating pediatric and non-elderly adult populations, we accepted cut-offs in the range 14-20, preferring the one available that was closest to 20. For separating non-elderly adults from elderly, we accepted cut-offs in the range of 54-70, preferring the one available that was closest to 60. Available seroprevalence data on more granular age strata were merged within the three main age groups.

### Data synthesis

We calculated for each eligible study, ratios of seroprevalence across children/adolescents, non-elderly adults, and elderly adults. The main analysis focused on the ratio of seroprevalence in the elderly versus non-elderly (non-elderly adults or any non-elderly, if pediatric and adult population data were not offered separately). In sensitivity analyses, we examined the ratios of seroprevalence in the elderly versus any non-elderly, and elderly versus strictly non-elderly adults. These ratios are “shielding ratios”^4^ and allow to evaluate whether elderly individuals (a high-risk group) were more protected (and if so, by how much) and if there were consistent patterns across different countries.

The observed ratios thus provide estimates of the extent of precision shielding achieved in different countries^4^ under the assumption that selection biases in sampling, test performance, and seroreversion are not substantially different in different age strata. Calculations were performed using the crude numbers (tested positive/total tested) in each age stratum; when these were not available, we used the adjusted seroprevalence estimates and converted the adjusted proportion to an equivalent number of seropositives. When both crude numbers and adjusted estimates were available, we examined whether the latter changed the results.

In secondary analysis, we examined the ratio of seroprevalence in children/adolescents versus non-elderly adults – to evaluate whether there was preferential shielding of pediatric populations.

We had anticipated that substantial heterogeneity may exist across countries to preclude formal data synthesis by meta-analysis. Therefore, we express results by using medians and also by describing studies with extreme values. We also formally estimated the between-study heterogeneity of these ratios using the I^2^ statistic^19^. In exploratory analyses, we evaluated whether results differed in high-income countries versus other countries (assuming that perhaps focused protection might be more feasible in the former).

## RESULTS

### Eligible studies

On February 8, 2022, SeroTracker had 547 entries of seroprevalence estimates which were described as national. After in-depth screening (Figure 1; excluded studies in Supplementary Table 2), 38 eligible studies were included in the analyses: 36 had separate seroprevalence data on an elderly age stratum, while the other two (Afghanistan, Oman) could only separate pediatric versus adult population seroprevalence.

**Figure 1.**
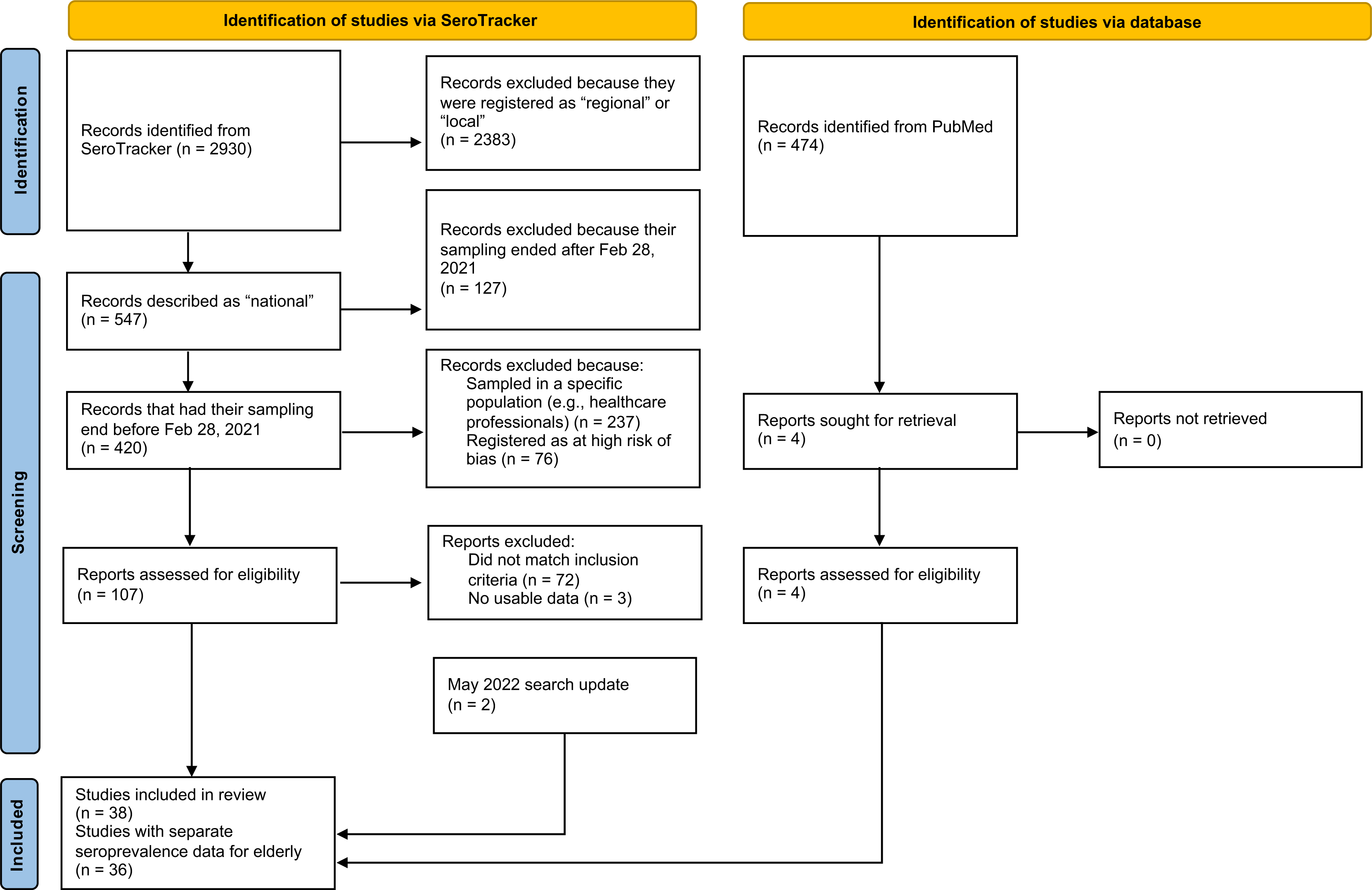
Flow chart for screening and selection of eligible studies. May 2022 search update: Among 3412 identified SeroTracker records, 9 reports were manually assessed for eligibility (after applying relevant SeroTracker filters according to first search), and 7 excluded. Details on the totally 79 reports excluded among 116 reports manually assessed for eligibility are in Supplementary Table 2.

### Study characteristics

Table 1 presents the 38 eligible studies. They came from 36 different countries (France and USA had two eligible studies each). More than half of the studies were performed in Europe (n=20), 13 were performed in Asia, 4 in the Americas and 1 in Africa. 25 of the 38 studies came from high income countries. Sample sizes varied substantially but tended to be higher in high-income countries. 24/38 studies had a total sample exceeding 5000, but this applied to only 6/13 studies from non-high income countries. 26/38 studies provided separate data for a pediatric population with cut-off ages varying between 14 and 19 years, and 36/38 provided separate data for an elderly population with cut-offs varying between 54 and 70 years. 11 studies assessed all antibodies, 7 assessed IgG and IgM, and 20 assessed only IgG. 20/38 studies performed all their sampling before or up to October 2020.

**Table 1.**
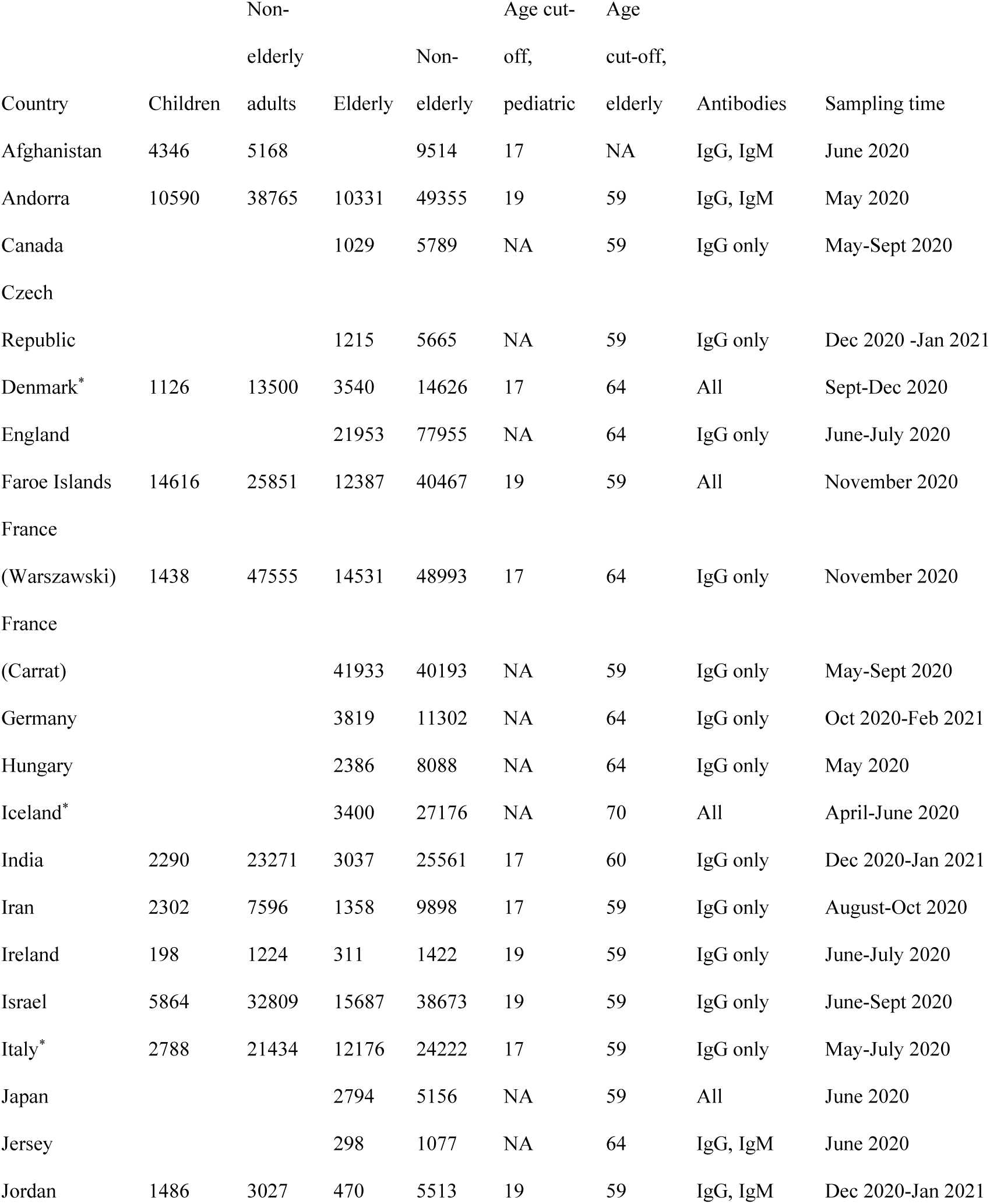
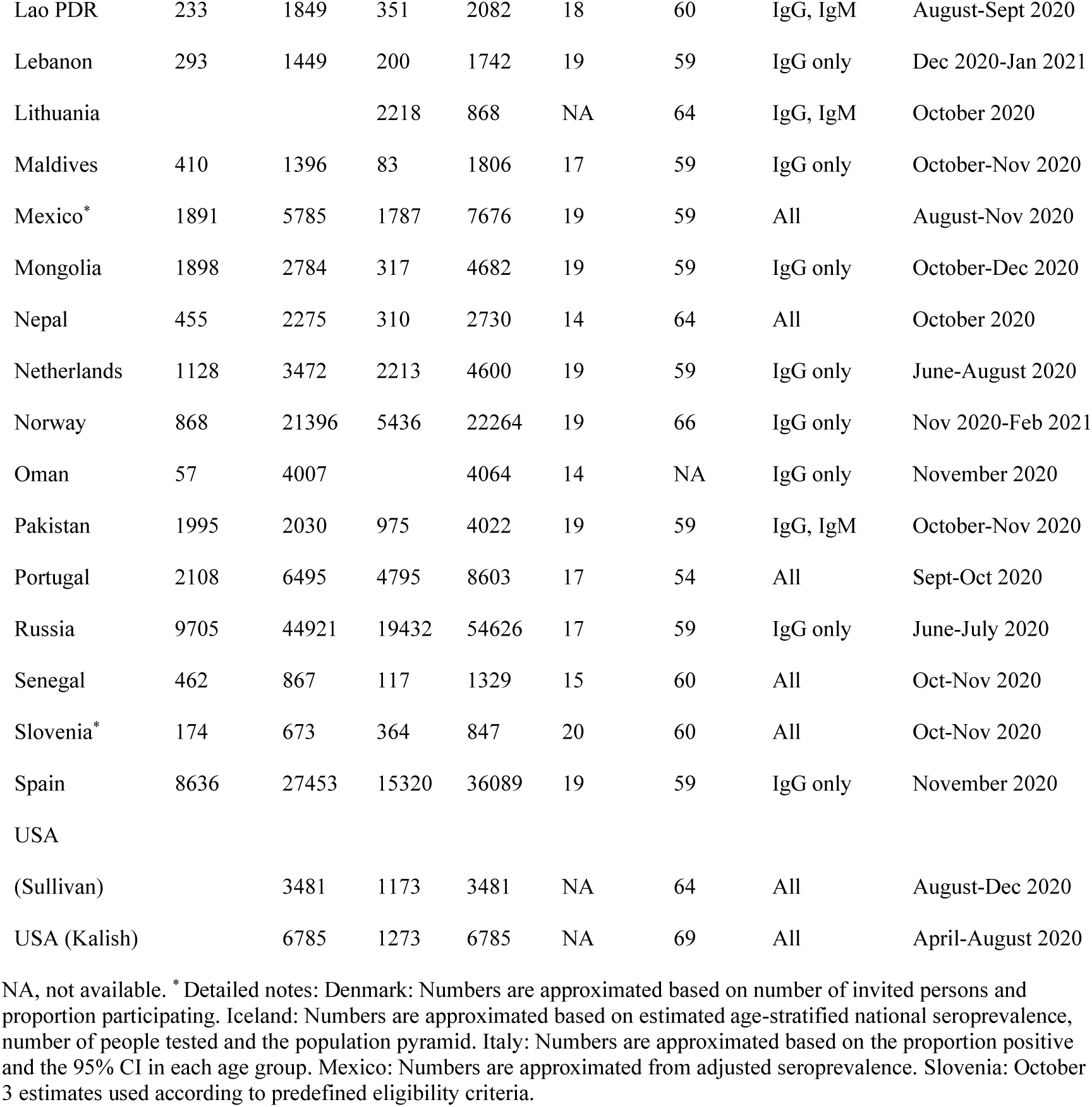
Eligible studies: population and sampling details

### Seroprevalence in the different age groups

Table 2 shows the seroprevalence estimates for the pre-specified groups of children, non-elderly adults, non-elderly, and elderly. There was a wide range of values from 0% in Faroe Islands to over 40% in the Czech Republic. Whenever available, adjusted seroprevalence estimates tended to be similar to unadjusted estimates with few exceptions (Table 2). Parameters used for adjustments are shown in Supplementary Table 3.

**Table 2.** Seroprevalence estimates (%) for age groups: unadjusted (and adjusted in parenthesis)

| Country | Seroprevalence<br>in children | Seroprevalence<br>in non-elderly<br>adults | Seroprevalence in<br>non-elderly | Seroprevalence<br>in elderly |
| --- | --- | --- | --- | --- |
| Afghanistan | 25.3 | 35.2 | 30.7 |  |
| Andorra | 12.6 | 11.0 | 11.3 | 13.1 |
| Canada |  |  | 2.1 | 0.7 |
| Czech Republic |  |  | 43.7 | 41.6 |
| Denmark | 6.5 (6.5) | 4.2 (4.3) | 4.4 (4.5) | 2.3 (2.3) |
| England |  |  | 6.1 (6.8) | 3.6 (3.2) |
| Faroe Islands | 0.0 | 0.0 | 0.0 | 0.0 |
| France (Warszawski) | 8.9 (9.8) | 6.7 (6.4) | 6.8 (6.5) | 4.2 (4.2) |
| France (Carrat) |  |  | 6.8 | 2.3 |
| Germany |  |  | 1.3 (1.9) | 1.1 (0.6) |
| Hungary |  |  | 0.6 (0.7) | 0.8 (0.8) |
| Iceland |  |  | 1.0 | 0.5 |
| India | 27.7 (27.0) | 25.6 (24.0) | 25.7 (24.5) | 28.2 (26.0) |
| Iran | (11.5) | (12.6) | (12.4) | (19.4) |
| Ireland | 1.5 (1.4) | 2.0 (1.7) | 1.9 (1.7) | 1.9 (1.7) |
| Israel | (7.2) | (4.0) | (4.5) | (2.2) |
| Italy | 2.2 (2.2) | 2.5 (2.5) | 2.5 (2.5) | 2.6 (2.6) |
| Japan |  |  | 0.1 | 0.1 |
| Jersey |  |  | 3.8 | 5.4 |
| Jordan | 36.2 | 33.8 | 34.6 | 34.5 |
| Lao PDR | 3.9 (4.2) | 4.9 (5.1) | 4.8 (5.0) | 8.8 (9.3) |
| Lebanon | 15.4 (17.8) | 15.9 (18.3) | 15.8 (18.2) | 18.5 (21.4) |
| Lithuania |  |  | 1.4 | 2.1 |
| Maldives | 4.9 | 15.2 | 12.8 | 31.3 |
| Mexico | 22.5 (22.5) | 27.8 (27.9) | 26.5 (25.7) | 18.6 (18.6) |
| Mongolia | 1.1 (0.8) | 1.7 (1.3) | 1.5 (1.1) | 1.3 (1.2) |
| Nepal | (8.8) | (15.7) | (14.1) | (10.7) |
| Netherlands | 2.4 (3.7) | 5.2 (4.9) | 4.5 (4.2) | 5.0 (4.9) |
| Norway | 1.8 (1.9) | 0.9 (0.8) | 0.9 (0.9) | 0.6 (0.5) |
| Oman | (23.5) | (21.5) | (21.5) |  |
| Pakistan | 4.2 | 8.9 | 6.6 | 8.7 |
| Portugal | (2.4) | (2.3) | (2.3) | (1.9) |
| Russia* | 21.6 | 15.6 | 16.6 | 17.4 |
| Senegal | 19.3 (19.3) | 31.7 (32.1) | 27.4 (27.4) | 24.8 (25.1) |
| Slovenia* | 4.9 | 5.6 | 5.5 | 2.1 |
| Spain | 7.8 (7.6) | 10.4 (10.5) | 9.8 (9.3) | 10.4 (10.3) |
| USA (Sullivan) |  | 5.6 (5.5) | 5.6 (5.5) | 2.9 (1.9) |
| USA (Kalish) |  | 3.8 (4.1) | 3.8 (4.1) | 3.6 (3.5) |

\* Detailed notes: Russia: Median percentage across tested regions. Slovenia: October 3 estimates used according to predefined eligibility criteria. Inverse-variance fixed effects meta-analysis used to combine age sub-strata.

### Ratio of seroprevalence in different age groups

Figure 2 shows the ratio of seroprevalence in the elderly versus the seroprevalence in non-elderly (non-elderly adults or non-elderly in general, if pediatric and adult population data were not offered separately). As shown (and as anticipated) there was large between-study variability with I^2^=98%.

**Figure 2.**
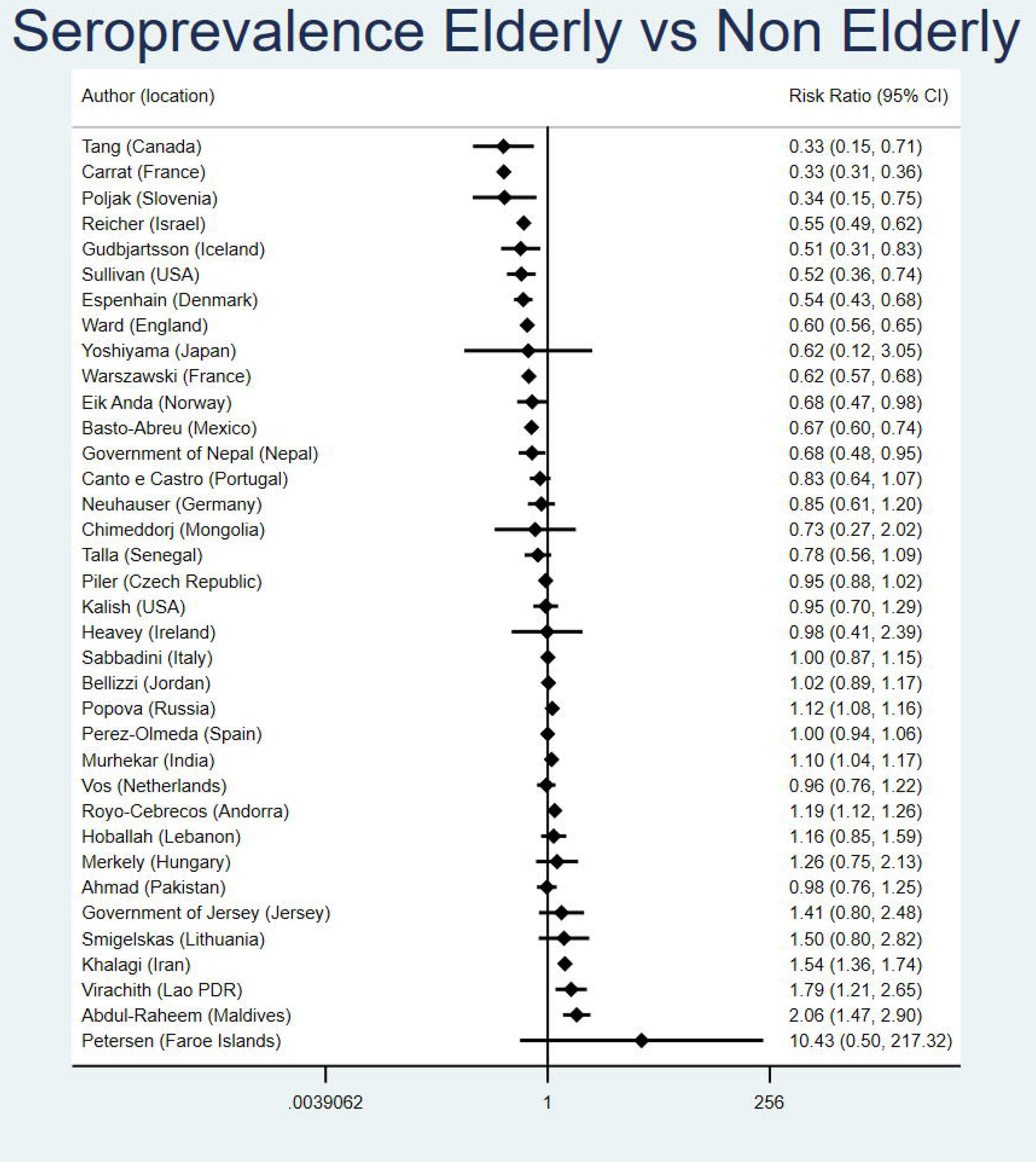
Seroprevalence ratio for elderly versus non-elderly (non-elderly adults or non-elderly in general, if pediatric and adult population data were not offered separately). See methods for definition of age groups. The presented 95% confidence intervals are estimated using crude counts in a 2 by 2 table for each study ([number elderly positive/number elderly tested]/[number non-elderly positive/number non-elderly tested]). When only adjusted seroprevalence estimates were available without crude data, these were converted to equivalent of number positive (number positive = adjusted seroprevalence x number tested in the specific age group).

The median ratio was 0.95 (0.90 if adjusted seroprevalence estimates were given priority in the calculations), suggesting a slightly lower seroprevalence in elderly populations and 23/36 studies had point estimates in this direction. For the two countries with two studies each, the point estimates were in the same direction, but the magnitude of the estimated protection of the elderly varied. Twelve studies with suggested protection of elderly and 6 studies with suggested inverse protection (higher seroprevalence in the elderly) had 95% confidence intervals excluding a ratio of 1.00. Canada, Slovenia, one of the two studies in France and (in adjusted analyses only) Germany and one of the USA studies suggested protection over 2.5-fold (ratio <0.40) with 95% confidence intervals excluding 1.00. Inverse protection of such magnitude (ratio >2.5) with 95% confidence intervals excluding 1.00 was not seen in any study.

Sensitivity analyses gave similar results: the median ratio of seroprevalence in the elderly versus any non-elderly was 0.95 with 20/36 studies offering point estimates in the direction of some protection of the elderly; the median ratio of seroprevalence in the elderly versus strictly non-elderly adults was 0.98 with 14/24 studies offering point estimates in the direction of some protection in the elderly.

In the comparison of pediatric populations versus non-elderly adults (Figure 3), there was again large between-study heterogeneity (I^2^=96%). The median ratio of seroprevalence was 0.89 and 15/26 studies presented point estimates in the direction of greater protection of children/adolescents than non-elderly adults. 15 studies had 95% confidence intervals excluding a ratio of 1.00 (with lower seroprevalence in the pediatric populations in 8 and higher in 7). Only one study (Maldives) showed a ratio of <0.40 with 95% confidence intervals excluding 1.00 and none had a ratio >2.5 with 95% confidence intervals excluding 1.00.

**Figure 3.**
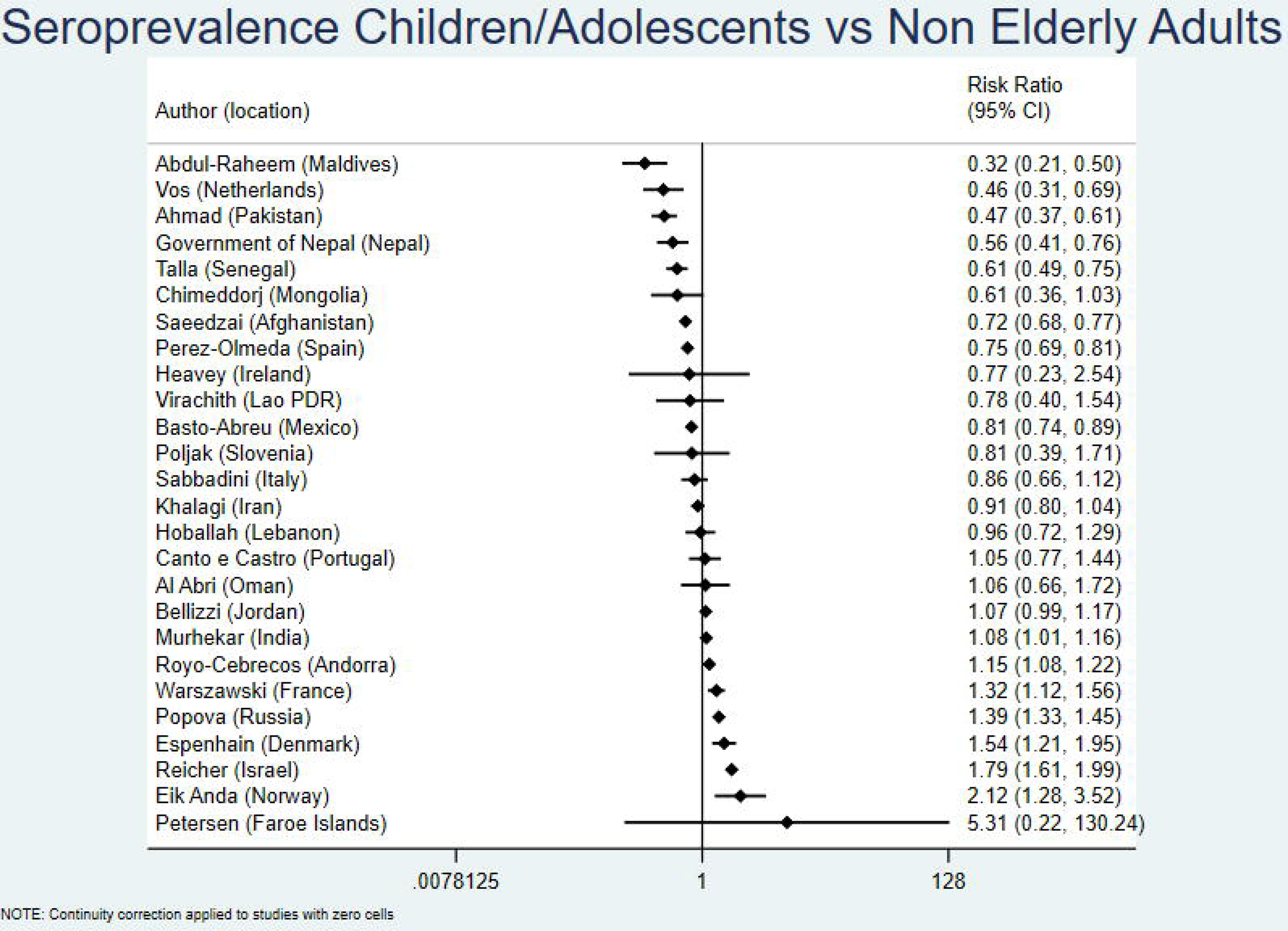
Seroprevalence ratio for pediatric populations versus non-elderly adults. See methods for definition of age groups. The presented 95% confidence intervals are estimated using crude counts in a 2 by 2 table for each study ([number pediatric population positive/number pediatric population tested]/[number non-elderly adults positive/number non-elderly adults tested]). When only adjusted seroprevalence estimates were available without crude data, these were converted to equivalent of number positive (number positive = adjusted seroprevalence x number tested in the specific age group).

### High income versus non-high income countries

For the main analysis of elderly versus non-elderly (non-elderly adults or non-elderly in general, if pediatric and adult population data were not offered separately), the median ratio was 0.85 in 25 studies done in high-income countries (0.83 if priority were given to adjusted estimates) and 1.02 in 11 studies done in non-high income countries. All 5 statistically significant estimates of >2.5-fold protection of the elderly were in high income countries. For a more modest protection threshold, all 9 estimates of >1.5-fold protection of the elderly (ratio <0.67) with 95% confidence intervals excluding 1.00 were in high income countries, and both of the 2 estimates of >1.5-fold inverse protection (higher seroprevalence in the elderly) with 95% confidence intervals excluding 1.00 were in non-high income countries.

## DISCUSSION

Our analysis of data from 38 national seroprevalence studies for COVID-19 showed that before the advent of massive vaccination there was large heterogeneity across countries on the extent to which elderly people in the community were protected or not from infection compared with younger populations. On average, there was very little extra shielding of the elderly. However, several countries apparently did achieve substantial precision shielding of this vulnerable group. Conversely, in a few countries elderly were apparently infected even modestly more frequently than non-elderly adults. Conclusive evidence for substantial preferential protection of the elderly in the community was seen only in some high income countries. In non-high income countries, the average ratio of seroprevalence between age groups suggested no preferential protection by age. There was also little difference in seroprevalence in children versus non-elderly adults overall, but the pattern differed across countries. On average, children were slightly less frequently infected than non-elderly adults.

These data suggest that precision shielding of vulnerable elderly populations is feasible, but strong shielding of the community-dwelling elderly populations was uncommon during the COVID-19 pandemic. It is unclear if this failure reflects practical difficulties of achieving major precision shielding, especially in disadvantaged settings^20,21^, or the fact that pandemic response policies may not have focused much on this aspect, instead aiming for more horizontal measures. Country-level responses may have differed in this regard, and even within countries heterogeneity may have existed across states and local communities. Given that a large share of COVID-19 deaths among community-dwelling people happen in the elderly, protection shielding exceeding 2.5-fold, as documented for 5 countries in our analysis, reflects roughly halving the total COVID-19 deaths among community-dwelling populations. Therefore, the benefit can be very large. Unfortunately, however, the countries that apparently did achieve some substantial shielding of their community-dwelling elderly, failed in protecting resident of long-term care facilities^7-13^, where infection fatality rates can be much higher (even 10-fold higher) than in community-dwelling elderly^17^. This resulted in numerous deaths of elderly residents in countries like USA, Canada, France, Germany, and Slovenia. Seroprevalence studies have documented extremely high rates of infection in nursing homes, much higher than in the community, in diverse countries, especially during 2020^9-13^.

Some caveats are worth discussing. First, while we used stringent criteria to select least biased studies, bias may still exist. Participants in serosurveys may differ from non-participants in infection risk.^15^ However, here this would be a problem only if the selection bias varied in different age groups.

Second, the probability of seroconversion after infection and the rapidity of seroreversion may vary depending on age.^22,23^ If anything, old age (and more symptomatic disease) tend to be associated with longer persistence of antibodies.^22^ If so, the precision shielding of elderly may have been slightly larger than what we calculate. Given that most studies evaluated here were done early in the pandemic, seroreversion was probably not large.

Third, for some studies, seroprevalence rates were very low and 95% CIs very wide. Depending on what adjustments are made, seroprevalence ratios might also differ in such cases, although in most studies we saw similar results for adjusted and unadjusted calculations.

Fourth, the counterfactual seroprevalence ratios in the absence of any restrictive measures are unknown. Evidence from influenza seroprevalence assessments suggests that often children and/or young adults may be infected more frequently than elderly individuals, perhaps due to greater mobility and exposures, but this is not absolute and may vary per year and location^24-27^. Extrapolations to SARS-CoV-2 are tenuous.

Finally, focused protection may have varied in subsequent phases of the pandemic with infection rate ratios across age groups. Vaccine availability in 2021 was typically prioritized for the elderly leading to shifts in the age distribution of COVID-19 impact.^28^ Vaccination also allowed more mobility and higher population exposure. After the Delta and Omicron waves, the vast majority of people were infected at least once in most countries.^29^ Even if precision shielding of the elderly can be achieved (as our data suggest), it is unknown whether it can be maintained effectively for pandemic-long circles lasting 2 or more years. Moreover, adverse consequences of trying to diminish exposures of vulnerable elderly may be substantial for their social well-being and their mental health^30,31^. Adverse consequences are likely for all age groups, including for children after school closures^32^.

Acknowledging these caveats, our analysis indicates that precision shielding was feasible in several high-income countries in the first year of the pandemic. However, most countries had no major differences in infection rates across age groups. Precision shielding or lack thereof may have substantially affected the eventual death toll and excess deaths^33^. These observations may be useful for future pandemic preparedness, especially for pathogens exhibiting large fatality rate variability across different population groups.

## Data Availability

All data produced in the present work are contained in the manuscript

---

**Supplementary Table 1:** Amendments/clarifications to protocol.

| Manuscript section | Amendment/Clarification | Rationale |
| --- | --- | --- |
| Eligibility for this subproject | For separating pediatric and non-elderly adult populations, we accepted cut-offs in the range 14-20, preferring the one available that was closest to 20. For separating non-elderly adults from elderly, we accepted cut-offs in the range of 54-70, preferring the one available that was closest to 60. | Clarification/specification needed for lower acceptable cut-off of pediatric group; for elderly cut-off accepted also one study (Portugal) with cut-off at 54 years (its exclusion gives similar results). |
| Analysis | The main analysis focused on the ratio of seroprevalence in the elderly versus non-elderly (non-elderly adults or non-elderly in general, if pediatric and adult population data were not offered separately). In sensitivity analyses, we examined the ratios of seroprevalence in the elderly versus any non-elderly, and elderly versus strictly non-elderly adults. | Clarification/specification needed |
| Analysis | Calculations were performed using the crude numbers (tested positive/tested) in each age stratum; when these were not available, we used the adjusted seroprevalence estimates and converted the adjusted proportion to an equivalent number of seropositives. When both crude numbers and adjusted estimates were available, we examined whether the latter changed results. | Clarification/specification needed; analyses were run with both crude and adjusted results and results are similar (as shown in the paper) |
| Analysis | The presented 95% confidence intervals for seroprevalence ratios are estimated using crude counts in a 2 by 2 table for each study. When only adjusted seroprevalence estimates were available without crude data, these were converted to equivalent of number positive (number positive = adjusted seroprevalence x number tested in the specific age group). | Clarification/specification needed |

**Supplementary Table 2.**
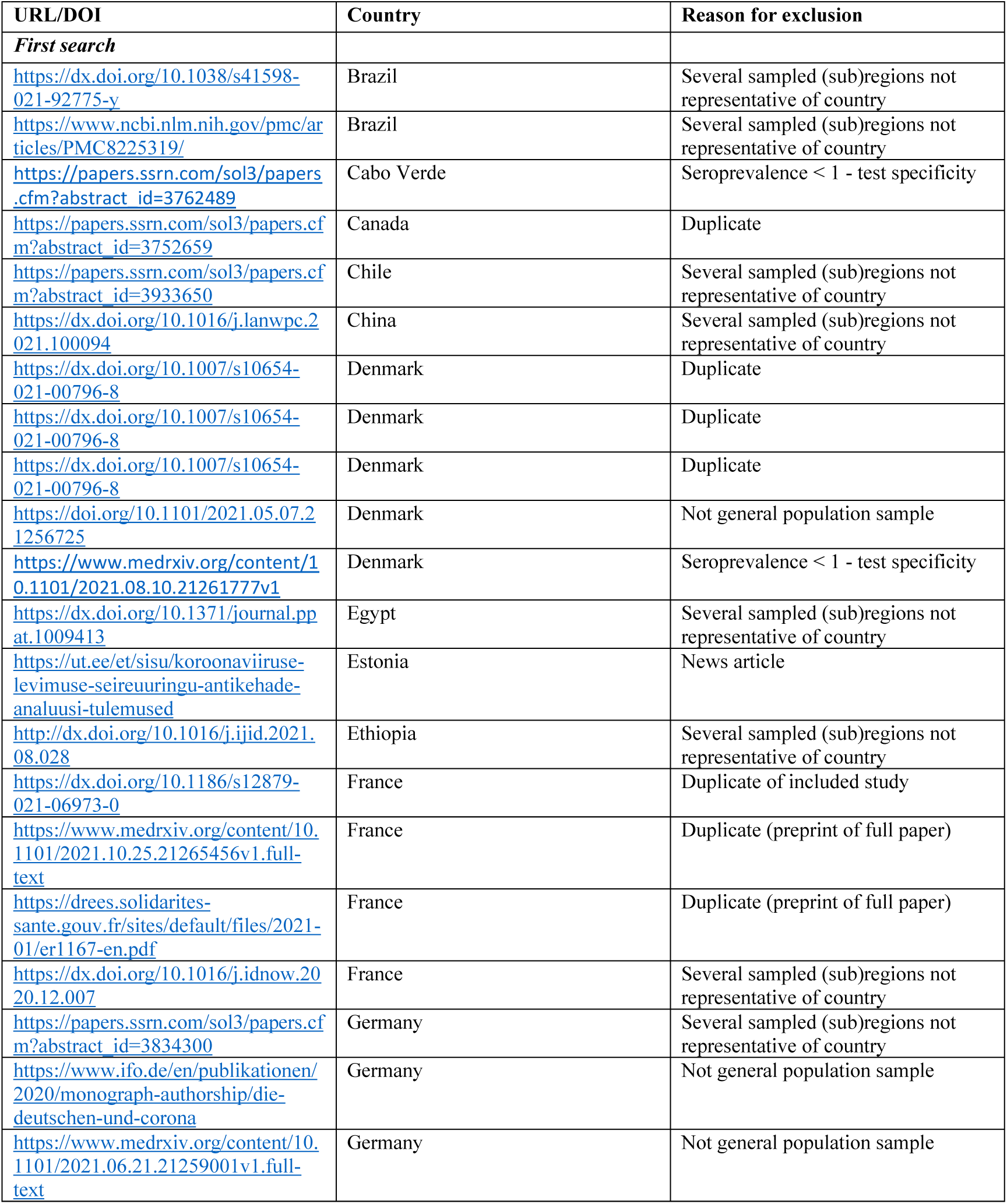
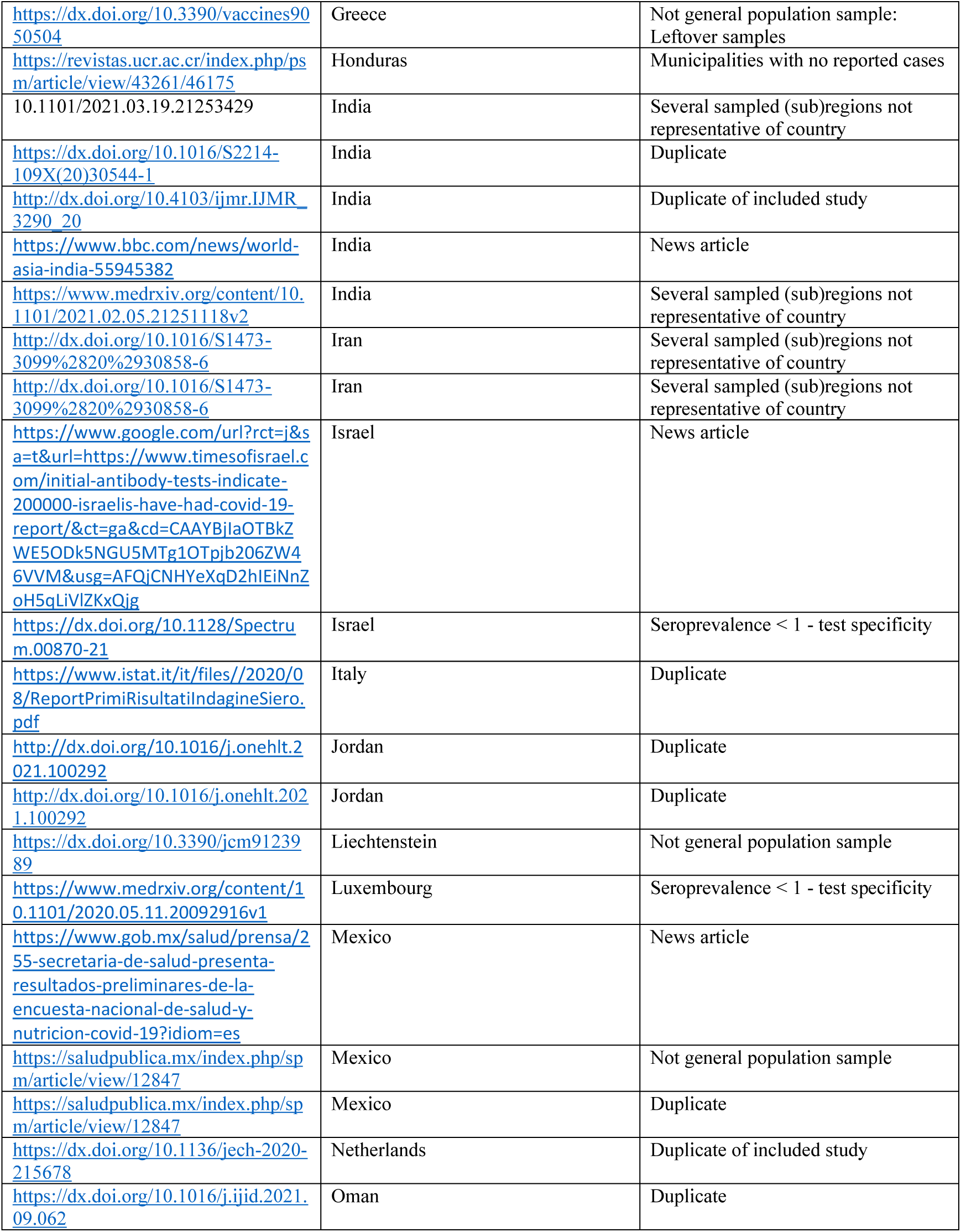
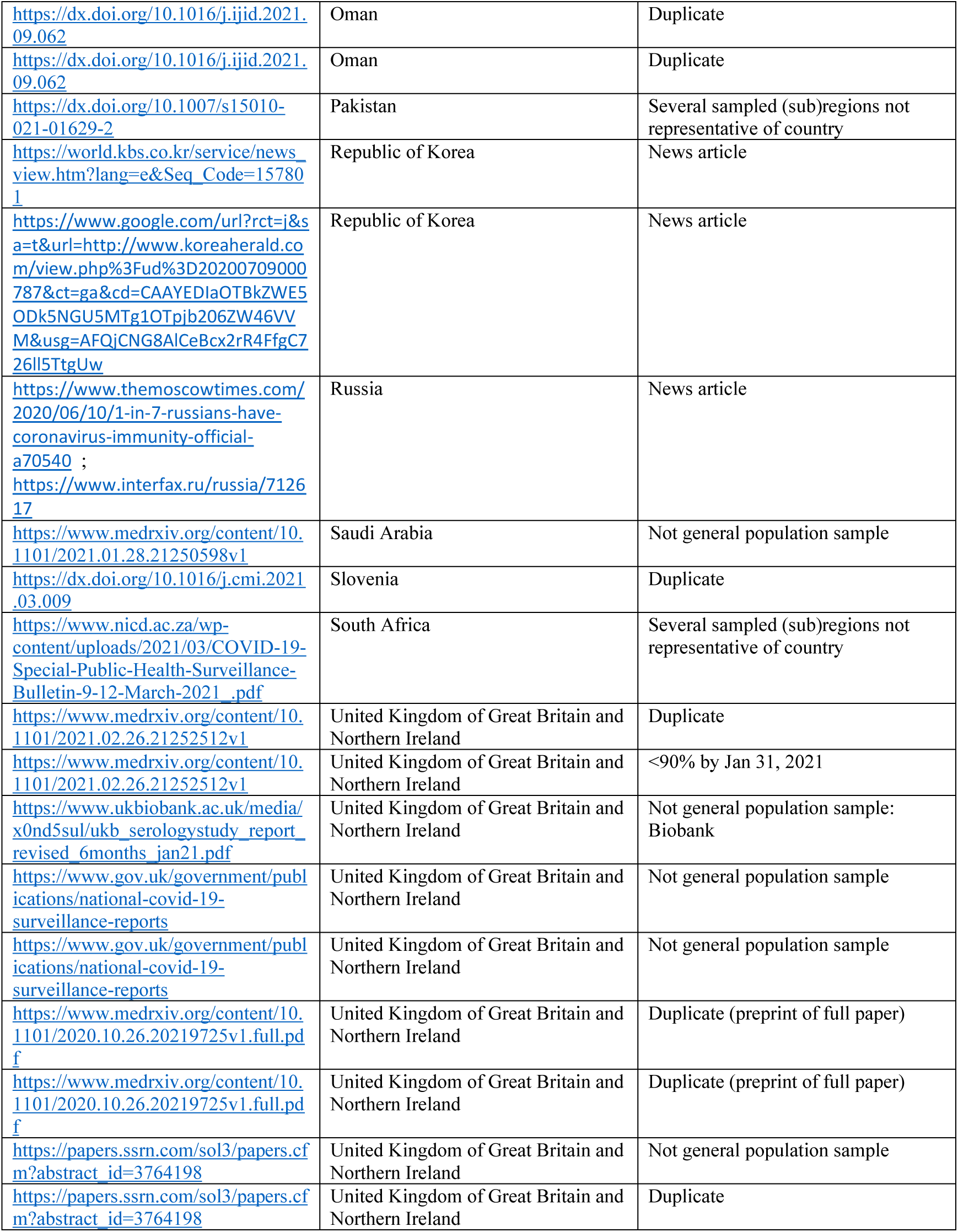
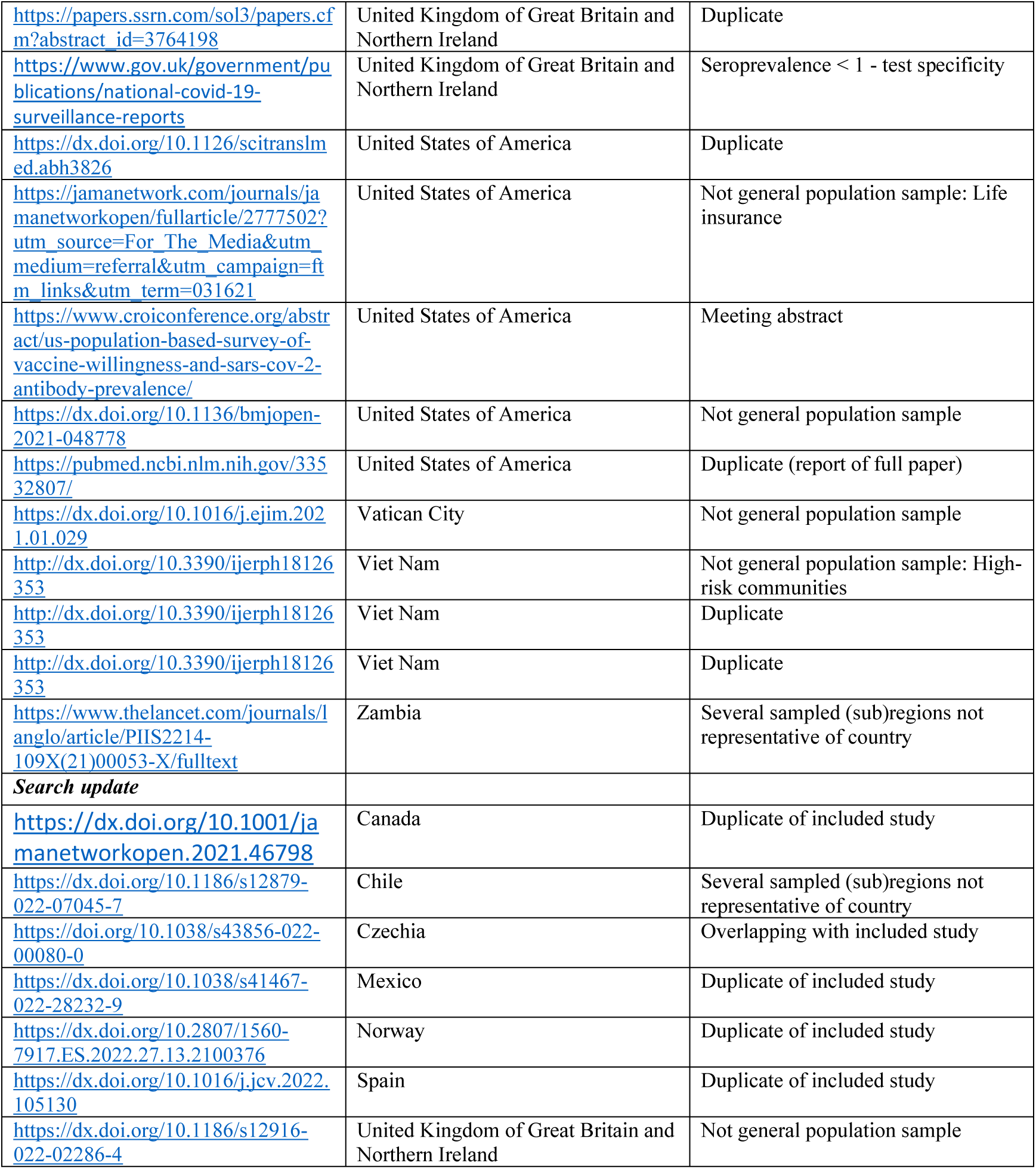
Details of excluded studies among reports manually assessed for eligibility (first search: 107 assessed, 72 excluded; search update: 9 assessed, 7 excluded).

**Supplementary Table 3:** Adjustments used in the adjusted seroprevalence estimates

| Country | Adjustments made |
| --- | --- |
| Afghanistan | NA |
| Andorra | NA |
| Canada | NA |
| Czech Republic | NA |
| Denmark | Test sensitivity and specificity using the Rogan-Gladen estimator |
| England | Test performance, and weighted to account for sample design and for variation in response rate (age, sex, ethnicity, region and deprivation) to be representative of the England population over 18 years |
| Faroe Islands | NA |
| France<br>(Warszewski) | Sample design, non-response, census calibration |
| France (Carrat) | NA |
| Germany | Non-response, test performance and seroreversion |
| Hungary | Design weighted. Response sample calibrated to known population counts by region, sex, and age categories. |
| Iceland | NA |
| India | Cluster sampling, sensitivity, specificity |
| Iran | Measurement error of the laboratory test, non-response bias and sampling design. |
| Ireland | Weighted to adjust for varying response rates in age-sex strata |
| Israel | Age, sex, time period, RT-PCR status, municipal strata, sampling |
| Italy | Non-response, region, age, sex, working status, province |
| Japan | NA |
| Jersey | NA |
| Jordan | NA |
| Lao PDR | Weighted for complex survey sample design, age and sex |
| Lebanon | Sex, age and area of residence, test performance |
| Lithuania | NA |
| Maldives | NA |
| Mexico | Test performance, and used sampling weights to adjust for selection probabilities and non-response rates (with post-stratification on region, sex and age group) |
| Mongolia | Test sensitivity and weighted prevalence rates according to age, sex, and provincial or district population size |
| Nepal | Survey design weights, age |
| Netherlands | Adjusted for survey design, weighted to match the distribution of the general Dutch population (based on sex, age, ethnic background, and degree of urbanization) and controlled for test characteristics |
| Norway | Rake weighting for population estimates of seroprevalence by age, sex, place of birth and county based on individual-level data for the invited sample (participants and non-responders) together with the corresponding distributions from the source population, provided by the Norwegian Population Register. Applied propensity scores for nonresponse adjustment and jackknife replicate weights for the raking procedure. Estimates subsequently corrected for test performance |
| Oman | Age group, sex, nationality |
| Pakistan | NA |
| Portugal | Adjusted for test performance, used sample weights and post-stratified by sex to adjust the seroprevalence extrapolating from the strata to the whole population |
| Russia | NA |
| Senegal | Weighted according to the age and sex distribution of the general population and adjusted test performance |
| Slovenia | NA |
| Spain | Characteristics of the random subsample of the fourth round |
| USA (Sullivan) | Test performance, design weights |
| USA (Kalish) | Age, region, sex, urban/rural, race, Hispanic, BRFSS survey response, sensitivity, specificity |

NA, not available

